# Measurement of doctor wellbeing prior to the Covid pandemic: a methodological systematic review

**DOI:** 10.1101/2024.10.03.24314759

**Authors:** G Simons, D Opalinski, J Jenkins, E Boxley, DS Baldwin

**Affiliations:** Centre for Workforce Wellbeing, University of Southampton, University Department of Psychiatry, College Keep, 4-12 Terminus Terrace, Southampton, SO14 3DT, United Kingdom; Clinical and Experimental Sciences, Faculty of Medicine, University of Southampton, Hampshire, United Kingdom; Solent NHS Trust; St Mary’s Hospital, Imperial College Healthcare, NHS Trust; St Georges University Hospitals NHS Foundation Trust; University Department of Psychiatry and Mental Health, University of Cape Town, South Africa

**Keywords:** Wellbeing, Measurement, Doctors

## Abstract

**Objectives:** Attention has been focused on health professional during and after the Covid-19 pandemic, but relatively little is known about wellbeing before the pandemic struck. We therefore wised to describe which wellbeing outcomes had been measured in doctors and which wellbeing outcome measurement instruments had been used with doctors, prior to 2020.

**Design:** A methodological review of existing literature.

**Setting:** MEDLINE, EMBASE, Cochrane Central Register of Controlled Trials (CENTRAL), PsycINFO, and the International Bibliography of Social Science were searched for all study types, in all languages.

**Outcome measures:** Wellbeing outcomes were categorised as being defined/operationalised in the aims, the methods or the results,**Error! Reference source not found**. and by whether the outcome used to represent wellbeing included the word wellbeing, another positive concept, a pathological symptom, a pathology, and were work-specific or doctor-specific. The outcome measurement instruments used were then categorised as published or unpublished and the frequency of use was collected.

**Results:** A total of 218 studies were included in this review. The majority of studies were not interventional (83.9%). The total number of unique outcomes used to capture wellbeing in the eligible studies was 57, with 369 non-unique outcomes. The percentage of outcomes used that contained the word wellbeing, its components and other positive concepts, was 69.9% (258/369). The percentage of negative concept use such as negative work context outcomes, symptoms of pathologies, or pathologies, was 30.1% (111/369). For the outcome “general wellbeing” alone, 92 different measurement tools were used. The Maslach Burnout Inventory was the most frequently used measurement tool for all outcomes, used in 16.3% of studies.

**Conclusions:** Wellbeing has been measured heterogeneously in doctors in terms of the outcomes and the outcome measurement instruments used. In approximately one-third of the times it was measured, the best that could be achieved was an absence of pathological symptoms, as a negative concept was used to operationalise it.

**Strengths and limitations of this study:** Strengths

- This methodological review includes 218 studies on doctor wellbeing
- This study utilised a novel methodology for determining eligibility and identifying outcomes for poorly defined concepts, such as wellbeing that reduces reviewer bias

Limitations

- This study relates only to the methods used in studies of doctor wellbeing
- The findings are based on studies published prior to the Covid-19 pandemic
- It was not possible to double extract all data

## Background

‘Wellbeing’ has no international consensus definition^1^ and inconsistent approaches to defining wellbeing in the literature have led to use of a diverse range of outcomes^2,3^. A systematic review of wellbeing measurement scales identified 60 different subjective wellbeing measurement tools^4^ and a more recent systematic review identified 99 measures^5^. In the UK, policy documents on doctor wellbeing, from the British Medical Association (BMA)^6-8^, Society of Occupational Medicine^9^, General Medical Council (GMC)^10^ and Health Education England^11^ that describe and make recommendations on doctors’ wellbeing, share the same lack of operationalisation of wellbeing. In a systematic review, only 11 of the 78 included papers contained an explicit definition of doctor ‘wellness’ ^12^.

There are multiple factors to consider in the measurement of wellbeing. These include why it is being measured: as examples, for screening of pathology; describing epidemiology; or assessing the efficacy of an intervention. Who measures also matters -an independent, impartial, third-party doing the measuring could be verified and is ‘objective’. However, subjective measurement is used most often for wellbeing, where answers can be ranked by the individual with numbers to make ordinal data. The locale of the measurement of wellbeing is relevant; it could be measured nationally (as examples, the NHS Staff Survey, BMA surveys), regionally or locally (for example, NHS Trusts). The context of measurement can be at work only, or life in general. It can be measured quantitatively or quantitatively depending on the context. It could be measured in terms of individual wellbeing currently or over a set or undefined time period, and in terms of concepts or determinants.

Pathologies are often used to describe wellbeing; in the systematic review of doctor wellness ‘burnout’ was the most common outcome measured^13^. This is understandable given the lack of a consensus definition and the tradition for pathology to be studied and described in medicine, but hinders progress on what wellbeing is, how it should be measured, and supported.

These factors emphasise the need for a careful analysis and synthesis of findings into a comprehensive summary of how wellbeing measurement in doctors has been undertaken. To provide a knowledge base on which a Delphi consensus can be built to address these issues, a systematic review is normally undertaken to identify possible outcomes and outcome measurement instruments^5^ and that approach is undertaken here.

### Research questions

1. Which wellbeing outcomes have been measured in doctors?
2. Which wellbeing outcome measurement instruments have been used with doctors?

### Methodology

No existing systematic review answered the research questions, so a protocol to answer them was developed and registered with Prospero (<u>Prospero ID: CRD42020141866</u>). This Systematic Review followed the Preferred Reporting Items for Systematic Review and Meta-Analysis (PRISMA) Checklist^14^.

### Eligibility criteria

The PICO (Participant, Intervention, Comparison, Outcome) method of identifying key concepts was utilised and adapted to the research questions.

**PICO Concepts used in the Systematic Review**

**Participants:** All grades and specialities of doctor

**Intervention:** For the research questions posed in this review no intervention needed to be captured, but the measurement of wellbeing was the process that needed to be captured

**Comparison:** No control or comparative measure was required

**Outcome:** Wellbeing was the outcome of interest

The definition of wellbeing utilised was ‘*Wellbeing is a state of positive feelings and meeting full potential in the world. It can be measured subjectively and objectively, using a salutogenic approach*’_15_. This definition was used as it was important that this review should capture objective as well as subjective, hedonist and eudemonic outcomes. It was also important for the synthesis and discussion of the results that measures could be grouped; into those that measure wellbeing, those that measure pathologies and negative outcomes, and those measuring positive concepts other than wellbeing. No language restrictions were placed. All types of study were included if they measured, or discussed measurement of, doctors’ wellbeing, for any purpose, including reviews and opinion pieces: including qualitative and quantitative measures, and measures which have been recommended but not yet utilised.

## Information sources

The following databases were searched with no restrictions.

### Bibliographic databases

1. MEDLINE,
2. EMBASE,
3. Cochrane Central Register of Controlled Trials (CENTRAL) Subject-specific databases:
4. PsycINFO,
5. International Bibliography of Social Science

### Search strategy

The search strategy (online supplemental file 1) was applied with no defined time period or language on 25/11/2019 (the first reported cluster of probable Covid-19 was reported in December 2019).

### Selection process

Titles and abstracts were assessed to allow irrelevant reports to be excluded by two researchers independently. When there was a disagreement about eligibility this was arbitrated by a third reviewer. Full-text versions of the potentially relevant reports were sought to assess their eligibility further. Papers that met the criteria had data extracted using a standardised form that allowed the planned outcomes to be captured.

### Data collection process

Data was extracted, collated, and assessed in Microsoft Excel^16^ using a data extraction form designed prior to extraction. If needed, authors were contacted for any missing data.

## Data items

The context of the studies:

- study type
- year of publication
- country conducted in

The sample studied:

- average age
- gender proportions
- specialities studied
- grades studied

### Study bias

- number approached
- number responded
- number at follow up

### The mechanism of measurement of wellbeing

- wellbeing in the title
- operationalisation of wellbeing, how it was described as an outcome (as an aim, in the methods or in the resultsError! Reference source not found.)
- delivery method for surveys
- outcomes used to capture wellbeing
- measurement instruments used to capture outcomes

### Risk of bias assessment

To reduce study risk of bias, results from running the search strategies in the bibliographic databases were exported and merged in Endnote^17^. Duplicate records of the same report were identified using the Endnote ‘Find duplicates’ function (year, title, volume, issue, pages) and by manual searching and removed. Multiple reports of the same study were linked. Two reviewers screened the studies and used the search function in Endnote Click^17^ to identify where wellbeing was operationalised.

## Synthesis of results

Due to the varied methodology, interventions and outcome measures used, a narrative synthesis was conducted. The operationalisation of wellbeing was conceptualised in three categories:

1) Studies that listed wellbeing as an explicit outcome in the results section,
2) Studies that listed wellbeing as an explicit outcome in the methods section (but did not use the word wellbeing in the results),
3) Studies that listed wellbeing as a measurement aim in the introduction or background (with no explicit mention of wellbeing in the methods or results).

Outcomes and outcome measurement instruments were identified using the criteria shown:

- Outcomes and measurement instruments that captured wellbeing, as defined by this study
- Outcomes and measurement tools that did not do so

There were many outcomes and measurement instruments that did not measure wellbeing but other positive concepts. They were either general or specific to the context of being at work, or being a doctor, and therefore the conceptual model of context was used:

- General positive concepts
- Work-specific concepts
- Doctor-specific concepts

The outcomes and measurement instruments that were not constructed to capture positive concepts were further categorised into:

- Pathologies
- Symptoms of pathologies

Outcome measurement instruments were categorised as above and as:

- Published
- Unpublished
- Quantitative
- Qualitative

### Reporting bias assessment

An update search at the end of the review process was not undertaken as this was a snapshot methodological review to capture measurement before the introduction of wellbeing roles within NHS Trusts as recommended by the NHS Staff and Learners’ Mental wellbeing commission and before Covid ^18^.

### Certainty assessment

The use of this method of categorising the operationalisation of a poorly defined concept allowed the reviewers to capture all the ways wellbeing had been measured. This methodology removed the bias of pre-defining what outcomes and/or measurement tools would be included when it was the methodology that was being studied.

To estimate the sensitivity of the search strategy: the percentage of relevant reports found out of the total in existence, the total number of relevant reports identified through database searches was divided by the total number of relevant reports identified through database searches and relevant systematic review backward citation searches. A total of 199 of the 235 relevant studies (84.7%) were identified by the search strategy.

The precision of the search strategy: the percentage of results found that were relevant out of all the results found was calculated by dividing the total number of relevant reports identified through database searches by the total number of irrelevant reports found through both systematic and backward citation searches. It was calculated that 4% of search results were relevant.

## Results

A total of 218 studies were included in this review. The studies that were eligible were heterogeneous in the contexts in which they studied doctors, the outcomes and outcome measurement instruments used.

## Study selection

### Study characteristics and contexts

The 218 studies identified were heterogenous, and the growth of publications on doctor wellbeing since 1985 was exponential. Studies that measured wellbeing to demonstrate the efficacy of an intervention (See Table 1) comprised 16.1% (n=35) of the studies, with 72% (n=157) capturing the epidemiology of wellbeing in doctors. Of these, 28.7% (n=45) did so at more than one time-point, and 71.3% (n=112) at only one time-point. The USA was the origin of the most studies (38.9%), and 99% of studies were conducted in Western cultures. In all the studies, 48.9% were undertaken in mixed speciality doctor populations and 61.5% in mixed grades of doctor. Those studies that captured the age of participants had a pooled median age of 38.9 years. The median percentage of male doctors in study populations was 53.5%.

**Figure 1.**
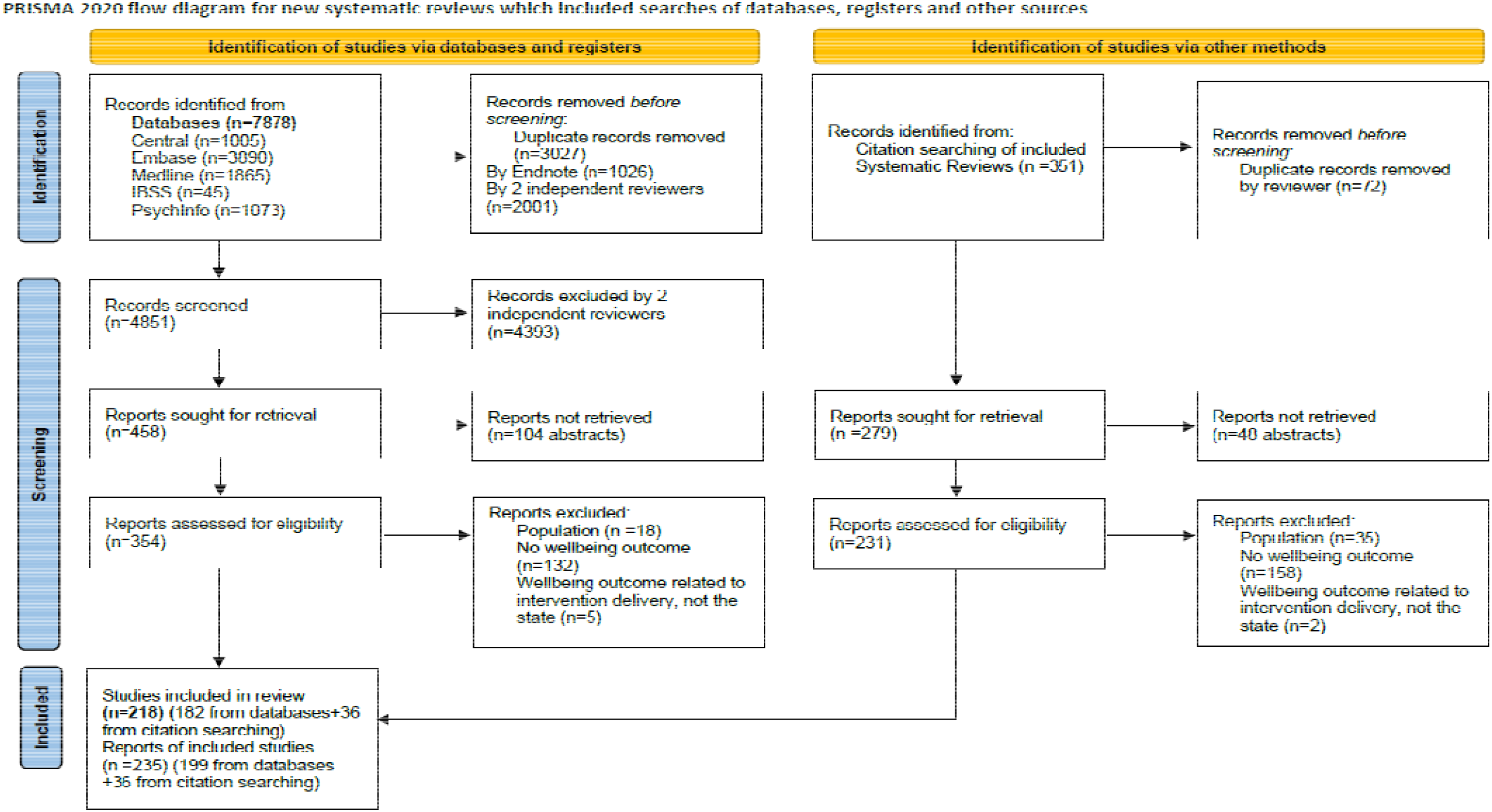
PRISMA 2020 flow diagram for this systematic review^19^(CC BY 4.0)

**Table 1.** Summary of interventional study findings.

| **Total number of studies**<br>**n=218 (%)** | **Wellbeing operationalised in results total**<br>**n=142** | **Wellbeing operationalised in methods total**<br>**n=49** | **Wellbeing operationalised in methods total**<br>**n=27** |
| --- | --- | --- | --- |
| **Interventional studies**<br>**35 (16.1% of all studies)** | **Wellbeing operationalised in results**<br>**n=16 interventional** | **Wellbeing operationalised in methods**<br>**n=16 interventional** | **Wellbeing operationalised in aims**<br>**n=3 interventional** |
| **Primary interventions (22.9% of interventional studies)** | Duty hour policy<sup>20</sup> | Duty hours<sup>21,22</sup> |  |
|  | Rest breaks<sup>23</sup> | Rota design<sup>24</sup> |  |
|  | Treadmill desks<sup>25</sup> |  |  |
|  | Site improvement plan implementation<sup>26</sup> | Operating Theatre noise<sup>27</sup> |  |
| **Secondary interventions (77.1% of interventional studies)** | Mindfulness<sup>28-32</sup> | Mindfulness training<sup>33-36</sup> | Mindfulness<sup>37,38</sup> |
|  | Solution focused seminars<sup>39</sup> | Cognitive Behavioural Therapy<sup>40,41</sup> | Cognitive behavioural therapy<sup>42</sup> |
|  | Dialogue groups<sup>43,44</sup> | Debriefing sessions<sup>45</sup> |  |
|  | Peer Mentoring<sup>46</sup> | Health and wellbeing workshops<sup>47</sup> |  |
|  | Reflection Rounds<sup>48</sup> | Meditation Training<sup>49,50</sup> |  |
|  | Management training<sup>51</sup> | Stress management training<sup>52</sup> |  |
|  | Resident Assessment Facilitation Team meetings<sup>53</sup> | Relaxation CDs<sup>54</sup> |  |

The interventions that were examined by studies were mainly secondary interventions (77.1%), i.e., group interventions that aim to strengthen the individual, rather than primary interventions that aim to strengthen the wider system. Mindfulness was the most studied secondary intervention ^7,55-64^, studied in 31.4% of interventional studies.

**Table 2.**
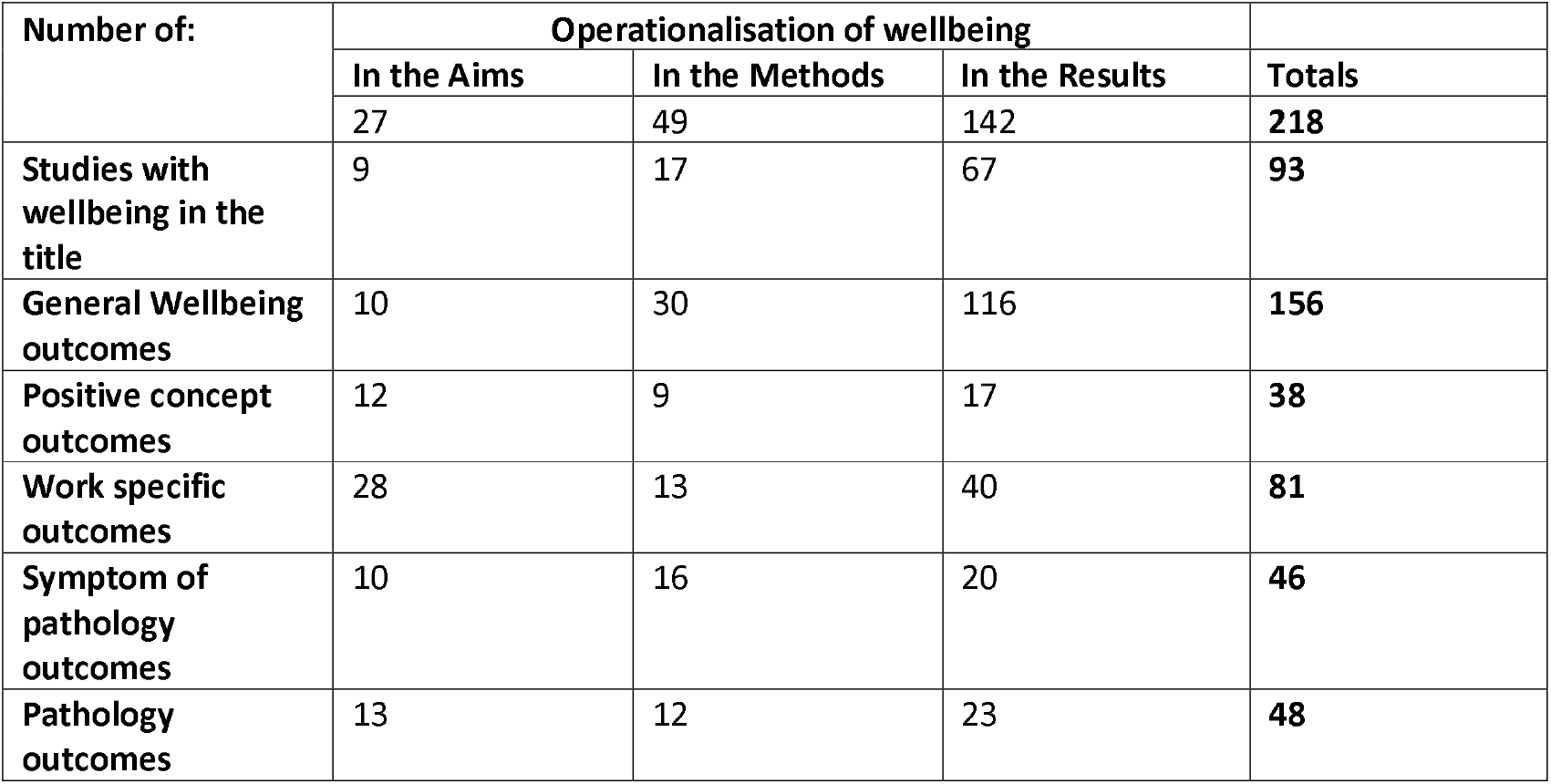
Summary of findings table, the way in which wellbeing was operationalised and measured. Studies could capture more than one outcome.

## Risk of bias in studies

### Selection bias

The majority (51.4%) of eligible studies were cross-sectional surveys capturing epidemiological data at a single time-point (71.3%), making the majority of studies unsuitable to investigate causality, and being at risk of sampling bias and conformity bias. This would have been mitigated by 72.7% of studies being conducted in multiple centres; however, most multi-centre studies were undertaken in hospitals in the USA, through the same healthcare provider group. Response rates for the cross-sectional surveys could be established in 41 of 112 studies (36.7%): the median response rate across studies was 62.7% (range 1.1-99%). Of the 16% of studies that were interventional 62.9% used a control and 42.9% used randomisation.

### Performance bias

All the interventional studies were unable to ‘blind’ participants, as doctors would be aware of what all the interventions comprised.

### Detection bias

Given the knowledge of doctors and their professional status there is potential for them to answer self-report outcomes more ‘correctly’ than the general population, to conform with social desirability. This would have been more problematic in the non-anonymous outcome collection methods.

### Attrition bias

For the prospective observational studies, cohort studies, randomised controlled trials, non-randomised controlled studies, and interventional studies with no control, the percentage of participants lost to follow up could be calculated in 31 of 80 studies (38.8%): the median attrition rate was 27.1% (range 0-55.1%).

### Reporting biases

Where results were missing, the authors were not contacted as this methodological review aimed to describe what was published about study design. Some interpretation was required to identify which measurement tool was chosen to capture the outcome wellbeing, usually involving a process of elimination as other outcome concepts were better defined and their measurement instruments could be paired with them more easily.

## Results of synthesis

### Research Question 1 – Outcomes measured in doctors

The total number of unique outcomes used to capture wellbeing in all the eligible studies was 57. Unique outcomes were those that represented a novel concept. Non-unique outcomes were those that described the same concept, using different wording. The total count of non-unique wellbeing outcomes used in the 218 studies was 369. The number and percentage of times an outcome was used that contained the word ‘wellbeing’ was 42.3% (156/369). The percentage of outcomes used that contained the word wellbeing, its components and other positive concepts, was 52.6% (194/369). If the positive work-context outcomes were also counted the positive outcome use percentage was 69.9% (258/369). The percentage of negative concept use such as negative work context outcomes, symptoms of pathologies, or pathologies, was 30.1% (111/369).

### Research question 2 -Wellbeing outcome measurement instruments used in doctors

For the outcome “general wellbeing” 92 different measurement tools were used, the most commonly employed being shown in Table 3. The measurement tools used could be classed as published wellbeing measurement tools (n=9), published measurement tools for positive concepts other than wellbeing (n=13), doctor-specific wellbeing measurement tools (n=10), work-specific measurement tools (n=9), measurement tools for symptoms of pathologies (n=6), pathology screening tools (n=13), unpublished study-specific measurement tools (n=20) and qualitative tools (n=12).

**Table 3.** The most commonly used published outcome measurement instruments to capture the outcome general wellbeing.

| Measurement Tool | Description of tool | Number of times used | References |
| --- | --- | --- | --- |
| Maslach Burnout Inventory | Published pathology screening tool | 8 | 66, 67 63,64,68,69 69 |
| WHO wellbeing index (5 item 5-point) | Published wellbeing measurement tool | 5 | 70-74 |
| 1 item VAS (100mm) | Published wellbeing measurement tool | 5 | 70,75-77 |
| LASA QOL | Published positive concept measurement tools | 4 | 78-81 82 |
| GHQ12 | Published pathology screening tools | 4 | 10 83-85 |
| Dupuy Psychological General Wellbeing scale | Published wellbeing measurement tool | 3 | 86-89 |
| Physician wellbeing index | Published Doctor-specific measurement tools | 3 | 9 90,91 |
| Stanford professional fulfilment (PFI) | Published Doctor-specific measurement tools | 3 | 92 93,94 |

The Maslach Burnout Inventory^65^ was the most frequently used measurement tool for all outcomes, used in 16.3% of studies; 22 studies that operationalised wellbeing as an explicit outcome in the results section, nine studies that operationalised wellbeing as an explicit outcome in the methods and three studies that stated wellbeing measurement was an explicit aim.

## Discussion

### Principal findings

The word ‘wellbeing’ was used in the title of 42.7% of studies, more so in studies that operationalised wellbeing in the results (72.1%). Importantly, each of the different ways of operationalising wellbeing included in this review (as an aim, or as an outcome in the methods, or in the results) was justified as each approach identified 10.5 -21.1% of the unique outcomes. In a systematic review of physician wellness^95^ 24 pre-defined ‘dimensions’ of wellness were used to categorise 75 non-unique outcomes in 43 studies published between 2010 and 2015. In comparison this review identified 57 unique wellbeing outcomes and 369 non-unique outcomes. Including studies that operationalised wellbeing as a measurement aim, or as an outcome in the methods, as well as in the results, allowed identification of all the ways that wellbeing in doctors has been measured. The heterogeneity of how doctor wellbeing was operationalised highlights both its multi-faceted nature and the need for a consensus approach to wellbeing research in doctors. A lack of operational definitions is not a problem unique to reviews of wellbeing research. A systematic review of core outcome set development studies found that no study defined how outcomes were differentiated and how final numbers of unique outcomes were determined ^96^.

The 92 measurement tools used to capture the outcome ‘General wellbeing’ alone again reflects the lack of a consensus operational definition for wellbeing, as does the finding that 21.7% of the measurement tools used were self-created. The finding that the Maslach Burnout Inventory, despite its associated cost, was the most commonly used tool (16.3% of all studies) shows a desire for a well operationalised concept and psychometrically tested tool. The use of a pathological outcome measurement instrument is worrying, as it limits the best a doctor can achieve to a lack of mental ill health. The finding that 30.1% of outcomes used to capture wellbeing were pathological or negative concepts reflects the ‘medicalisation’ of the concept of wellbeing, which is inherently positive and holistic.

### Strengths and weaknesses of the study

This systematic review included a large number of studies (n=218) compared to the previous systematic reviews identified through database searches ^9,97-104^ which identified a median of 28.5 eligible studies (range 3-81). The inclusion of all study types and languages, as well as searching diverse databases should have reduced selection bias in this systematic review. Reporting bias will be present, as published work is accessed more easily and it was not practically possible to search ‘grey literature’, given the 7878 studies retrieved in the existing database searches.

This systematic review utilised a novel way of categorising how a poorly defined concept – ‘wellbeing’ – is conceptualised in publications. It achieved this by coding whether the concept featured in the aims, method or results. This strategy enabled Adobe Acrobat Pro DC ^5^ and Endnote click ^105^ to be used to identify where the concept was mentioned, which reduced reviewer error and subjective bias as reviewers are only required to identify if the word is being used as an outcome.

The inductive collection of outcomes without a pre-conceived framework used in this systematic review has been used in other systematic review of wellbeing measurement^12,13,106,107^ and provides a more complete picture of the doctor wellbeing landscape by allowing for a comprehensive synthesis of all methods for measuring doctor wellbeing.

To mitigate selection bias, double screening was used with 10% of full-text articles being double-screened at the ‘Title and Abstract’ stage. Reviewers were not blind to authors and institutions, but these data did not need to be read to assess for eligibility; selection bias risk was therefore low. Ideally, all studies would have had data extracted by two independent researchers, but this was only possible for 36.5% of the reports. An audit was performed of 10% of the extracted outcome data for each of the 3 ways wellbeing could be operationalised: as an aim, in the methods or in the results, with no disagreements. It was not possible to comment on the sensitivity of this review compared to others, as the only other methodological systematic review in this area did not report the necessary data ^81^

### Meaning of the study and Implications for clinicians, policymakers and future research

The heterogeneity of outcomes used to measure doctor wellbeing hinders comparisons between studies looking at epidemiology or the efficacy of interventions. There is no consensus about which measurement tools are appropriate for the measurement of doctor wellbeing, leading to many different and often self-created tools being utilised. Meta analysis will not be possible until the same outcome measurement instruments start to be used.

### Conclusion

Wellbeing has been measured heterogeneously in doctors in terms of the outcomes and the outcome measurement instruments used. Just under a third of the times it was measured, the best that could be achieved was an absence of pathological symptoms, as a negative concept was used to operationalise it.

The results of this systematic review highlight the importance of Core Outcome Sets, in which a minimum agreed set of outcomes and recommended measurement tools to capture them allow comparisons across studies. The use of a Core Outcome Set, including recommended outcome measurement instruments would provide the optimal environment for synthesis and meta-analysis to occur in the field of doctor wellbeing.

## Data Availability

All data produced are available online at

http://dx.doi.org/10.5258/SOTON/D2318

## Author contribution

Simons G: Designed the study, collected, analysed and interpreted the data and wrote the manuscript.

Opalinski D: Collected, analysed and interpreted data and contributed to the manuscript. Jenkins J: Collected, analysed and interpreted data.

Boxley B: Collected, analysed and interpreted data.

Baldwin DS: Funding acquisition, overseeing design of the study, data collection and analysis. Edited and approved the manuscript.

## Acknowledgements

We thank Dr Aimee O’Neill and Professor Julia Sinclair for their contributions to discussions of notions of wellbeing and their connotations.

## Data sharing statement

All data supporting this study are openly available from the University of Southampton repository at https://doi.org/10.5258/SOTON/D1933

