## Supplemental File 1 for "Measurement of doctor wellbeing prior to the Covid pandemic: a methodological systematic review"

**Supplementary File 1 Systematic Review**

**EMBASE Search Strategy**

| Search Term number | Search Terms for EMBASE Ovid  Embase+Embase classic 1947-2019 |
| --- | --- |
| 1. (Participants)   Title and abstract | (doctor* OR physician* OR medical practitioner* OR clinician* OR surgeon* OR house officer* OR housemen OR foundation trainee* OR core trainee* OR speciality trainee* OR registrar* OR associate specialist* OR fellow* OR medical intern* OR medical resident* OR medical attending* OR consultant* OR general practitioner* OR hospitalist* OR internist* OR orthopedist* OR orthopaedist* OR urologist* OR anesthetist* OR anaesthetist* OR anesthesiologist* or anaesthesiologist* OR intensivist* OR obstetrician* OR gynecologist* OR gynaecologist* OR andrologist* OR cardiologist* OR pulmonologist* OR gastroenterologist* OR hepatologist* OR nephrologist* OR neurologist* OR epileptologist* OR dermatologist* OR endocrinologist* OR diabetologist* OR ophthalmologist* OR otolaryngologist* OR oncologist* OR radiologist* OR neonatologist* OR pediatrician* OR paediatrician* OR geriatrician* OR gerontologist* OR pathologist* OR hematologist* OR haematologist* OR phlebologist* OR immunologist* OR virologist* OR microbiologist* OR medical geneticist* OR psychiatrist* OR physiatrist* OR rheumatologist*).ab,ti. |
| 2. (Method)   Title and abstract | (measure*).ab,ti. |
| 3. (Outcome)   Title and abstract | (wellbeing or well being).ab,ti. |

**MEDLINE Search Strategy**

| Search Term number | Search Terms MEDLINE Ovid  Ovid Medline R 1946-2019 |
| --- | --- |
| 1. (Participants)   Title, abstract, keyword | (doctor* OR physician* OR medical practitioner* OR clinician* OR surgeon* OR house officer* OR housemen OR foundation trainee* OR core trainee* OR speciality trainee* OR registrar* OR associate specialist* OR fellow* OR medical intern* OR medical resident* OR medical attending* OR consultant* OR general practitioner* OR hospitalist* OR internist* OR orthopedist* OR orthopaedist* OR urologist* OR anesthetist* OR anaesthetist* OR anesthesiologist* or anaesthesiologist* OR intensivist* OR obstetrician* OR gynecologist* OR gynaecologist* OR andrologist* OR cardiologist* OR pulmonologist* OR gastroenterologist* OR hepatologist* OR nephrologist* OR neurologist* OR epileptologist* OR dermatologist* OR endocrinologist* OR diabetologist* OR ophthalmologist* OR otolaryngologist* OR oncologist* OR radiologist* OR neonatologist* OR pediatrician* OR paediatrician* OR geriatrician* OR gerontologist* OR pathologist* OR hematologist* OR haematologist* OR phlebologist* OR immunologist* OR virologist* OR microbiologist* OR medical geneticist* OR psychiatrist* OR physiatrist* OR rheumatologist).ab,ti |
| 2. (Method)   Title and abstract | (measure*).ab,ti. |
| 3. (Outcome)   Title and abstract | (wellbeing or well being).ab,ti. |

**CENTRAL Search Strategy**

| Search Term number | Search Terms for CENTRAL |
| --- | --- |
| 1. (Participants)   Title Abstract Keyword | doctor* OR physician* OR “medical practitioner*” OR clinician* OR surgeon* OR “house officer*” OR “housemen” OR “foundation trainee*” OR “core trainee*” OR “speciality trainee*” OR registrar* OR “associate specialist*” OR fellow* OR "medical intern*" OR "medical resident*" OR "medical attending*" OR consultant* OR “general practitioner*” OR hospitalist* OR internist* OR orthop?edist* OR urologist* OR an?esthetist* OR an?esthesiologist* OR intensivist* OR obstetrician* OR gyna?cologist* OR andrologist* OR cardiologist* OR pulmonologist* OR gastroenterologist* OR hepatologist* OR nephrologist* OR neurologist* OR epileptologist* OR dermatologist* OR endocrinologist* OR diabetologist* OR ophthalmologist* OR otolaryngologist* OR oncologist* OR radiologist* OR neonatologist* OR p?ediatrician* OR g?eriatrician* OR g?erontologist* OR pathologist* OR h?ematologist* OR phlebologist* OR immunologist* OR virologist* OR microbiologist* OR “medical geneticist*” OR psychiatrist* OR physiatrist* OR rheumatologist* |
| 2. (Method)   Title Abstract Keyword | Measure* |
| 3. (Outcome)   Title Abstract Keyword | wellbeing OR “well being” |
